# The Feasibility, Acceptability, and Perceived Need for Suicide Initiatives for Children in UK Primary Schools

**DOI:** 10.64898/2026.11.03.26367158

**Authors:** Barakh Abdul Salam, Pooja Saini, Molly McCarthy, Nina Smith, Emma Ashworth

## Abstract

**Background:** Currently, there are no specific suicide prevention programmes for primary (elementary) schools within the UK. However, given the rise in suicidal thoughts and/or behaviours amongst children under 11 years, the feasibility, acceptability, and perceived need for these programmes need to be assessed.

**Aims:** This qualitative scoping study aimed to explore whether there is a perceived need for suicide prevention programmes in primary schools and, if so, what these should look like.

**Sample(s) & Methods:** 92 participants took part in the study. Semi-structured interviews were conducted with mental health professionals (N= 22), school staff (N= 8), and parents of children who had experienced suicidal ideation, behaviours, or been bereaved by suicide (N=7). Focus groups were conducted with secondary school students (N= 36) and primary school students (N= 19) across 5 schools. Data were analysed using thematic framework analysis.

**Results:** Three main themes were identified: 1) the necessity of suicide prevention programmes for primary schools, 2) barriers affecting delivery and potential solutions, 3) the transition period from primary to secondary school.

**Conclusion:** Overall, participants supported the need and acceptability of suicide prevention programmes for primary schools, particularly due to the rising concerns about suicidality among younger children and evidence that suicide-related discussions are already occurring within peer groups. However, careful consideration needs to be given to ensure it is delivered in a safe and age-appropriate way, with a focus on dispelling myths, promoting help seeking, and increasing knowledge.

---

Within the United Kingdom (UK), there are no explicit suicide prevention programmes for children in primary (elementary) schools (aged 4-11 years), despite rates of suicidal ideation, behaviours, and attempts increasing in children and young people (CYP) (1,2). In 2023, 109 suicidal deaths were reported in 10- to 17-year-olds within England and Wales (3). Rates among younger children are increasing in the UK, with 25% of suicide deaths in CYP occurring in 10- to 14-year-olds (4). Growing numbers of primary school-aged children are experiencing mental health crises severe enough to result in Emergency Department attendance, with recent national data indicating a marked increase among 6-9-year-olds, raising important questions about the adequacy of early intervention and prevention within school (5). Thus, it may be necessary to begin prioritising preventative care at an earlier age.

There are multiple risk factors for CYP suicidal thoughts and attempts (6,7). Firstly, studies have reported that a quarter of 11- to 16-year-olds with a mental health condition have self-harmed or attempted suicide. However, accessing appropriate support remains difficult, with 270,300 CYP awaiting mental health support in 2025 (8,9), while many are denied access (10). Bereavement by parental suicide also increases the likelihood that CYP will experience poor mental health, suicide attempts, and self-harm (11–13). Indeed, the highest rates of self-harm have been identified in bereaved children under 10 years (14). However, Wardley (15) highlighted the absence of UK data on bereaved children, calling for national data collection to inform the planning and resourcing of appropriate support.

Another element affecting CYP is the transition from primary (3-11 years) to secondary school (11-16 years) (16). This is a significant developmental milestone and one of the most stressful events for CYP due to differences in environment, structure, and academic demands (17,18). Figures suggest that 31% of CYP report difficulties in adjusting to their new schools, and that students feel less safe and have reduced prosocial tendencies as they get further into the year (19), thus potentially increasing their risk of experiencing suicidal thoughts. The transition period is also when CYP begin accessing the Internet independently and more frequently (20,21), which may increase exposure to cyber bullying, and inappropriate and harmful content (22–24). For instance, Robinson et al. (25) found that 21.4% of children under 14 years were first exposed to suicide and self-harm content online, and that this exposure resulted in self-harm in 16.9% of participants.

Currently the PHSE National Curriculum includes teaching children how to take care of mental health, unpleasant feelings, and how to seek support (26). However, despite increasing evidence that primary school-aged children may be experiencing or exposed to suicide ideation, attempts, or bereavement, they do not currently have routine access to suicide prevention education to reduce myths and stigma or provide information to promote help-seeking. An increasing risk of suicidal ideation as children transition to secondary school suggests an upstream approach in primary schools may be beneficial, intervening early to equip CYP with help-seeking skills and prevent later suicidal behaviours/attempts. However, there is currently limited information on suicide prevention initiatives in primary schools, including whether there is a perceived need for them, whether they would be acceptable to deliver and, if so, what they would need to look like.

Conversely, there is evidence to suggest that suicide prevention programmes can be implemented safely and effectively within secondary schools. Schools are frequently viewed as a source of universal access for CYP, providing opportunity to incorporate early preventative programmes (27,28). Breaking the Silence and Signs of Suicide are two evidence-based programmes for secondary schools, which teach CYP how to spot the signs of suicide, respond, and signpost (29,30). A recent systematic review examined suicide prevention programmes for adolescents (aged 10 to 19 years) globally and found that 82% of studies reported significant positive effects on suicidal behaviours, 94% reported improvements in knowledge, and 67% reported improvements in help-seeking intentions or behaviours (31).

However, a commonly cited concern regarding suicide prevention education is that discussing suicide may be distressing, developmentally inappropriate, or could inadvertently introduce suicidal ideas (32). Similar concerns have been identified in relation to suicide-risk screening, although evidence to date does not support an iatrogenic effect, with one study finding repeated suicide-risk screening among children aged 8-12 was not associated with increased suicidal thoughts (33). Indeed, research suggests that well-designed, school-based suicide prevention programmes do not increase suicidal thoughts or behaviours and can improve help-seeking, mental health literacy, and stigma reduction when implemented sensitively and with family involvement (33–35). This is supported by the Government’s updated Keeping Children Safe in Education (KCSIE) Guidance, which now states that all staff must be aware of suicide and self-harm (36). Furthermore, delivering these programmes within schools could help address existing barriers to accessing mental health services and strengthen relationships between peers (37,38). Additionally, for low income, and racial and ethnic minority CYP who have mental health needs and are at risk of suicide, schools are especially crucial resources, given these populations are much more likely to receive support exclusively in educational settings (39,40). Nevertheless, parents and educators may have concerns regarding their own knowledge, confidence, and ability to respond appropriately to children’s questions or disclosures (41).

Limited information currently exists on the effects of discussing suicidality with CYP under 11, including whether it is safe or necessary. Although young children face multiple risk factors, suicide prevention programmes have yet to be encouraged. Hence, this study aimed to explore the feasibility, acceptability, and perceived need of a suicide prevention programme in UK primary schools. The scoping study specifically looked to explore (1) whether there is a perceived need for suicide prevention programmes in primary schools and, if so, what these should look like, and (2) whether primary school children want mental health interventions and what they would like to learn about.

## Methods

### Design

A qualitative design was utilised to explore the experiences of stakeholders across England. Specifically, semi-structured, one-to-one interviews were conducted with school staff (SS), mental health professionals (MHP), and parents (P) who had supported children with suicidality. Focus groups were conducted with primary school pupils (PSP) and secondary school pupils (SSP). Ethical approval was granted by [blinded] Research Ethics Committee (ref: [blinded]).

### Participants and Recruitment

Adult stakeholders and schools were recruited through voluntary/opportunity sampling via adverts distributed by email to existing networks of the authors and on networking apps (e.g., LinkedIn). Individuals interested in participating were provided with a participant information sheet and opt-in consent form. Gatekeepers recruited students from Years 5-6 in primary schools (aged 9-11 years), and Years 9-11 in secondary schools (aged 13-16 years). They were asked to select students of mixed ages, different sociodemographic backgrounds and experiences. Gatekeepers were asked to exclude students experiencing mental health difficulties or suicidality, or for whom participation might be distressing. Once students were selected by the school, verbal consent was taken from the students and parental information sheets and opt-in consent forms were sent out. Only students with parental consent could participate. Written assent was then taken from students at the beginning of the focus groups. Students were also provided with an information sheet prior to assent, using ‘child-friendly’ language and read aloud by the research assistant. Focus groups were conducted with 3-6 students per group (although one individual interview was conducted because no other parents/carers returned consent forms).

Thirty professionals participated (see Table 1). Twenty-two were mental health professionals (including charity workers, therapists, researchers) and eight were school staff members. Participants varied across age and gender, with the majority identifying as female (63%). While the majority participated in interviews, two focus groups were conducted within an organisation at the participants’ request; one included six participants and another three participants. Seven parents of children who had experienced suicidal ideation or behaviours, or who had been bereaved by suicide took part in one-to-one semi-structured interviews (see Table 2). 71.4% of parents identified as female, 28.6% were aged 35-44 and 71.4% were aged 45 to 54.

**Table 1.** Demographic characteristics of professionals and school staff.

| *Demographic characteristics* |  | *Total* | *Mental Health Professionals* | *School Staff* |
|---|---|---|---|---|
| Participant (*N*) |  | 30 | 22 | 8 |
| Age | 18 – 24 | 6.7% (*n*=2) | 4.5% (*n*=1) | 12.5% (*n*=1) |
|  | 25 – 34 | 6.7% (*n*=2) | 9.1% (*n*=2) | - |
|  | 35 – 44 | 23.3% (*n*=7) | 27.3% (*n*=6) | 12.5% (*n*=1) |
|  | 45 – 54 | 20% (*n*=6) | 22.7% (*n*=5) | 12.5% (*n*=1) |
|  | 55 – 64 | 16.7% (*n*=5) | 13.6% (*n*=3) | 25% (*n*=2) |
|  | Prefer not to say | 26.7% (*n*=8) | 22.7% (*n*=5) | 37.5% (*n*=3) |
| Gender | Male | 10% (*n*=3) | 13.6% (*n*=3) | - |
|  | Female | 63.3% (*n*=19) | 63.6% (*n*=14) | 62.5% (*n*=5) |
|  | Prefer not to say | 26.7% (*n*=8) | 22.7% (*n*=5) | 37.5% (*n*=3) |

**Table 2.** Demographic characteristics of parents.

| *Demographic characteristics* |  | *Parents* |
|---|---|---|
| Participant (*N*) |  | 7 |
| Age | 35 – 44 | 28.6%<br>(*n*=2) |
|  | 45 – 54 | 71.4%<br>(*n*=5) |
| Gender | Male | 28.6%<br>(*n*=2) |
|  | Female | 71.4%<br>(*n*=5) |

One primary school (19 students) and four secondary schools (36 students) took part (see Table 3). Primary school children participated in four mixed-gender focus groups. In secondary school 1, seven students took part, across one focus group and one interview (aged 13-15 years, of mixed gender).

**Table 3.** Demographic characteristics of student participants.

| *Demographic characteristics* | Total | Secondary School 1 | Secondary School 2 | Secondary School 3 | Secondary School 4 | Primary school 1 |
|---|---|---|---|---|---|---|
| Participant *(N)* | 55 | 12.7% (*n* =7) | 12.7% (*n* =7) | 23.6% (*n* =13) | 16.4% (*n* =9) | 34.5% (*n* =19) |
| Age range (*Years*) |  | 13 – 15 | 14 – 15 | 13 – 16 | 13 – 15 | 9 – 11 |

Secondary school 2 was an all-girls’ school, with seven students taking part, resulting in two focus groups (aged 14-15 years). In secondary school 3, three focus groups were conducted with 13 students aged 13-16 years (mixed gender). In secondary school 4, two focus groups were conducted with nine students (aged 13-15 years, mixed gender).

### Materials

Four semi-structured interview schedules were created for professionals, parents, primary students, and secondary students. These schedules guided discussions while allowing exploration of unexpected themes. Professional participants were asked about their experiences working with CYP in relation to mental health and suicidality, their perspective on the need for suicide prevention programmes, and their feedback on the resources that could be used to develop a programme. The parental interview schedule was similar, exploring whether they felt a need for suicide prevention in primary schools, why, and what it should involve. Secondary school students were asked about whether they thought it would be appropriate to have a suicide prevention programme in primary school and if so, what should or should not be included. Focus groups with primary students did not mention suicide to ensure safety and instead focused on what participants knew about mental health, what they would like to learn about, and how they would like to be taught.

### Procedure

All interviews with adult participants were completed online and recorded via Microsoft Teams. Interviews and focus groups with CYP were completed in their schools. At the beginning of the focus group, the researcher emphasised to all participants that they could stop the study at any time point, skip questions, and did not have to talk about any individual experiences. Participants were informed that there was no set suicide prevention programme that the researchers were trying to implement; this was highlighted so that participants felt comfortable to talk freely without feeling like there was a right or wrong answer.

All interviews and focus groups were conducted by the research assistant (BA). Focus groups were conducted in a private, quiet room to ensure confidentiality. After each session, participants received an age-appropriate support document signposting relevant services and were encouraged to ask questions. Interviews and focus groups lasted on average 37 minutes (8-66 minutes).

### Analysis

Transcripts were reviewed and edited by the research assistant to ensure accuracy and anonymity. Data were analysed using NVivo 15 using the five stages of thematic framework analysis (familiarisation, coding, creating themes, reviewing themes, and defining themes) (42). The researcher (BA) first familiarised themselves with the data and generated initial codes, before engaging in discussions with other members of the research team (EA, PS) to generate initial themes. Remaining transcripts were then coded against this framework, and themes were reviewed and defined. Final themes were agreed, defined, and interpreted by the research team.

### Findings

Three themes were identified: 1) the necessity of a suicide prevention programme for primary schools, 2) barriers affecting delivery and potential solutions, 3) the transition from primary to secondary school (Table 4).

**Table 4.** Themes and sub-themes.

| Themes | Sub-themes |
|---|---|
| Theme 1: The necessity of a suicide prevention programme for primary schools | Primary students experiencing suicide ideation |
|  | Hearing about suicide from peers |
|  | The role of the internet/ media |
|  | Importance of early intervention and lack of available services |
| Theme 2: Barriers affecting delivery and potential solutions | Parental concerns |
|  | Staff comfortability and knowledge |
|  | Keeping everyone fully informed |
| Theme 3: The transition period from primary to secondary school | Coping with changes |
|  | Future knowledge |

### The necessity of a suicide prevention programme for primary schools

#### Primary students experiencing suicide ideation

Mental health professionals and school staff discussed how suicidal thoughts and behaviours were common in primary schools and described working with primary-aged children who had struggled with suicidality: *“I was experiencing children who were disclosing suicidal ideation or thoughts of suicide, thoughts of ending their own life from around the age of seven, eight.”* (MHP7). They described children experiencing passive suicide ideation, often referring to wanting to be dead or not wanting to be here, rather than active ideation or plans: *“She would regularly say that she didn’t want to be here anymore or ‘wish I was dead,’ you know, and at the age of like 9 and 10”* (SS26).

Parents also mentioned that thoughts of suicide had started in primary school: *“we were kind of Year 4 [aged 7-8 years] where we were thinking about wanting to die”* (P3). For those whose difficulties started later, parents believed that prevention sessions earlier on in primary school could have helped their child before they reached a later crisis, equipping them with knowledge of how to seek help: *“and I think back to when they were 9 or 10, if they’d been having, you know, sort of awareness sessions in school around suicide at that age, I think it definitely could have helped”* (P1).

#### Hearing about suicide from peers

Many participants mentioned that primary-aged children were already aware of suicide, particularly because their peers were sharing suicidal thoughts with them, often before teachers were aware that there was a problem: *“I wouldn’t just be thinking about the children who experience those thoughts, but the children who are hearing from their peers that their peers experiencing that, because a lot of the time they hear it before we do”* (SS8). Both parents and professionals also mentioned that their primary-aged children have heard suicide being discussed by other children in school:

> *“Children know what suicide is. It’s being spoken about on the playground. And from my own personal experience of that… the amount of times that I had to contact my daughter’s primary school to say, ‘this child has spoken about suicide to my daughter’ And that’s a primary school age” (MHP9)*

Secondary school students also indicated that it was common to come across the word within primary school: *“they’re going to probably find out about it themselves, like I knew and I was like, what? 9 or 10, and no one told me”* (SSP3). Moreover, when discussing what should be included in a mental health session with primary school students, suicide was suggested by the students themselves: *“things like suicide and like bad thoughts”* (PSP2).

However, this was not the case for everyone. Some did think it might be too soon to introduce the word suicide to primary schools: *“it might be like more harm than good because, I mean like, exposing them to like what actually suicide might mean. Because like when I was in primary, I didn’t know what suicide meant*.” (SSP28). Some participants discussed the possibility of not “*directly bringing suicide up”* (MHP15), and instead using case studies and scenarios to discuss with the children, for example:

> *“We say he’s put a plastic bag over his head and then we talk about what steps do we take after that? And one of like the key things in it is about like exploring what that behaviour, what we’ve just seen, what does it mean? Like what was his intentions? So just exploring things that a little bit further I think is important as well.”* (MHP15)

#### The role of the Internet/media

Professionals felt that CYP were spending more time on social media than previous generations, which they partly attributed to parental concerns about children’s safety outside, with implications for social relationships and CYP’s mental health:

> *“I think another driver for that is social media, again, because I think as parents, we don’t feel that it’s safe for them to go out… It’s sad because, you know, we’re social animals we need that sort of human interaction, but their friendships are not as deep anymore because everything’s digital, so they, you know, they don’t hug each other or anything anymore. It’s sad.”* (MHP4)

Parents also described difficulties navigating the *“online world”,* which was referred to as a “*third parent”* (P2), as well as having a “*dehumanising effect”* (P5). Professionals highlighted how suicide, mental health, and self-harm are themes featured in many films and television series, which could be seen positively, by rising awareness, or negatively, by trivialising the topic. They felt this meant children are aware of what suicide is and, in some cases, may increase the cognitive availability of suicide for vulnerable CYP:

> *“Media portrayals, like certain TV series intend to raise awareness, but they can sometimes make suicide seem more common or less serious. And in some vulnerable young people, it could even increase harm because some people do take things the wrong way.”* (SS28)

CYP emphasised how necessary it is that children are taught about online safety and the dangers of social media, to ensure they are equipped with the skills needed to keep themselves safe: *“but if they didn’t learn it [online safety] from their parents, they’re going to learn it somewhere else. They’re not going to know what to do then.”* (SSP4). Professionals emphasised that by prohibiting CYP from accessing social media, they would do it without the parents’ knowledge, promoting unsafe use: *“I think that might be the risk. Parents worry that if they’re exposed to it [suicide], but a lot of the time these children are on social media and being exposed to so much anyway without parents overseeing it.”* (MHP14). To overcome this, participants felt that *“Internet safety”* (MHP22) should be included if a suicide prevention programme were to be implemented. Secondary school students also suggested that students should be taught about identifying “*truth and fake news”* (SSP12).

### Importance of early intervention and lack of available services

The need for suicide prevention programmes was emphasised by professionals due to the lack of available mental health services, with some CYP needing more support than schools could provide but not meeting the high thresholds for CAMHS support:

> *“CAMHS is exceptionally challenging for us to get in place, even when we know that that’s what we need. And then what we find is young people don’t reach the criteria, this threshold for CAMHS, which seems to get higher and higher and higher every single time I make a referral. But then we have something in my area, which is called [organisation name], which also kind of provide support for young people. But then if there’s real suicide ideation or action planning or whatnot, then they’re deemed too high for that [organisation name]. So, we’ve got these children that fall in this middle area.”* (SS8)

Another issue raised was the lack of accessible and effective support for young neurodivergent individuals, given that they are at increased risk of both mental health difficulties and suicide:

> *“My daughter is autistic and has ADHD. And so, I think for that group of children and young people, mental health services are particularly awful… they’re just not equipped to care for, in my opinion, children, young people who’ve got kind of additional needs over and above 6 sessions of CBT.”* (P3)

With neurodiversity, participants had mixed feelings on how to best support these individuals with sensitive sessions, with some suggesting that “*when it’s adapted for neurodivergent children*” (MHP4) it will be useful for all students and that the programme should be “*inclusive by nature*” (SS29). However, other participants believed sessions should differ for neurodivergent individuals as *“they don’t learn in the same way and don’t always take on the same message”* (SS25). Mental health professionals suggested that some neurodivergent individuals may need to “*have some separate teaching or some teaching revisited”* (MHP7).

### Barriers affecting delivery and potential solutions

#### Parental concerns

A common barrier highlighted to programme delivery was parental concerns, related to religious, cultural, personal beliefs, or “*because they don’t understand something or they fear it”* (MHP10): “*if you say we’re running this suicide prevention thing at primary school, that’s going to make the parents go ‘my kids are not doing that’”* (P4). Participants described how parents’ discomfort around the topic could lead them to refuse participation, as with other personal, health, or relationship education sessions:

> *“We always get big letters home about ‘In this session, we’re going to talk about sexual health or whatever. If you’ve got concerns’ and there are parents that will always contact the school with, ‘I don’t really want my child listening to this’.”* (P3)

It was also noted that the subjective experiences of parents can carryover onto the child, which in that case creates a bigger barrier: *“I think there might be some resistance from parents… I think sometimes parent anxiety feeds into child anxiety and then, you know, that might be an issue”* (MHP14).

#### Staff comfortability and knowledge

Another barrier was the comfortability of school staff in discussing suicidality, particularly given the lack of training and knowledge. Professional participants questioned whether they are equipped to deliver a programme on suicidality: *“‘am I the right person to deliver this?’”* (MHP5). Some participants also noted that “*members of staff who’ve dealt with that [suicidality]”* (SS27) may find this particularly difficult.

When discussing who should deliver a suicide prevention programme, opinions varied. Some believed that schoolteachers were unsuited due to *“very little support or supervision”* (MHP7) available for staff after delivering PSHE sessions. It was also highlighted that teachers “*might have certain views or values”* (MHP7) around suicide, potentially affecting delivery.

Students preferred an external professional delivering the potential programme, with a teacher present, believing this would benefit everyone and ensure the message was communicated appropriately: *“Like I’d rather them sit back and listen with the kids then and have someone else talk because they’ll have their own opinions and they might not express it properly to the children.”* (SSP6). This was also supported by primary-aged students, who expressed the importance of the students’ comfortability and how students will feel safe by having their teacher present: *“Kind of both because the classroom teacher, they’ve been with them longer. You get to feel more comfortable with the teacher, but with another person in, they might have different views that they can learn.”* (PSP1). However, some students believed that having the teacher deliver the session would be more effective as teachers tend to become a secure figure in a student’s life, therefore students *“won’t mind telling them if something wrong”* (SSP3).

#### Keeping everyone fully informed

All participants felt it was important for everyone to be fully informed. Participants described how understanding why the programme was being delivered would help teachers feel more comfortable around delivery: *“I think if teachers know the statistics and if teachers understand the research as to why we’re doing it, I think they would do it”* (MHP2). Similarly, informing parents that suicide is a current issue affecting CYP may prevent barriers: *“I think it’s being open and transparent to the parents early doors and sending out as much information as you possibly can and really highlighting exactly why you’re doing it”* (MHP4).

Most participants suggested workshops for parents and school staff before programme implementation, alongside involving them in its delivery, to alleviate concerns: *“Parents need to be brought on the journey. They need to understand why things are being done, the way that it’s going to be done, have the opportunity to be able to ask questions.”* (MHP 5)

Despite recognising the programme’s importance, participants also emphasised that CYP should have the option to not participate: *“Children who have got, you know, are struggling with mental health, they do definitely need an opt out”* (MHP30). Primary school students also emphasised they should “*have the choice*” (PSP1) but felt that it was important for students their age to have lessons on mental health: *“Yeah, give them the option to like do it because if they feel comfortable doing it, then I think you should learn about it”* (PSP13).

### The transition period from primary to secondary school

#### Coping with changes

CYP participants discussed the anxieties associated with the transition phase, particularly the unfamiliar environment, including *“new teachers and new people they don’t know”* (PSP9) and the fear of not getting *“into the same school as your friend”* (SSP25). They also discussed how this can be seen as both positive and negative: *“there’s a lot more people for you to be friends with, but then there’s also a lot more people for you to not be friends with and have arguments with.”* (SSP5). The increase in workload was another challenge during the transition phase, as well as the fact that school gets more serious: *“I think there’s like definitely more work in secondary school and they also in secondary school like tell you about more topics and they don’t like shield you from anything.”* (SSP2).

Ultimately, students described the transition as *“just a stressful experience in itself”* and *“nerve-wrecking”* (SSP5), highlighting its potential impact on emotional wellbeing, which could potentially lead to suicidal thoughts.

Besides the external changes, participants also highlighted that children experience puberty and hormonal changes, which can affect emotional wellbeing. They therefore emphasised the importance of teaching children to regulate heightened emotions and access support:

> *“You’ve got obviously then puberty may have come before, but maybe after. So, you’ve got all that kind of emotional change as well and, you know, physical changes happening in the body and, you know, they can evoke real big emotions. And I think that, you know, those big emotions need to be addressed.”* (SS30)

As a result, participants highlighted the importance of CYP knowing how to seek help and support peers, particularly in Year 6, to equip them before transitioning to secondary school: *“we need to equip them with those skills and knowledge to have that awareness, but also how to support themselves or how to support a friend.”* (MHP22). School staff also emphasised the value of preventative sessions, particularly around death in general and grief: *“Talking about, you know, dying and the impact of real-life, impact of how life carries on for other people and on the point of view of other people who are left behind”* (SS30).

Participants emphasised that the stigma surrounding suicide can hinder help-seeking and the ability to have open conversations. Therefore, participants stressed the need for equipping CYP to develop emotional literacy to be able to support one another: *“Even at the very lowest level, being able to be there for each other as friends, as kids together and being able to support one another through difficult times.”* (MHP24).

#### Future knowledge

Although participants placed strong emphasis on the struggles CYP face when moving to secondary school, not all students are negatively impacted. However, participants argued that all students should receive support and knowledge around mental health and suicide prevention, even if they do not need it at the time, as they might need it later, either for themselves or to support others: *“they might not think it’s that important, but I think when they get older, they’ll realise how important it is to have the lesson”* (PSP13). Early prevention was also perceived to have the potential to reduce suicidality: *“If we can get into that really early with people, then we’re not going to have the numbers of suicides that we’re having now in adults.”* (MHP9).

Parents were also identified as a barrier to children accessing support by not allowing them to receive “*useful information*” (P3). Participants linked this to suicide stigma and the contribution of generational attitudes, highlighting the importance of early preventative care to challenge these patterns. This was supported by primary students discussing mental health: *“when you are older and you have like kids of your own, they might need like help”* (PSP13).

## Discussion

This scoping study investigated the feasibility, acceptability, and perceived need for suicide prevention programmes in primary school. Specifically, the study aimed to explore primary school children’s views on mental health interventions, including what they would like to learn, and stakeholders’ views on the need for and content of suicide prevention programmes for primary-aged children.

The qualitative findings generally indicated a perceived need for suicide prevention programmes in primary schools, although there was hesitancy for some. Participants emphasised that many primary school-aged children experience suicidal ideation, and mental health professionals reported supporting CYP as young as seven years old. This is consistent with national data indicating 109 suicide deaths among 10- to 17-year-olds in 2023 (3) and with Miller et al.’s (43) findings that almost half (45.92%) of children aged 5-10 years reported passive or active suicidal ideation. Nevertheless, participants highlighted that children’s awareness and understanding of suicide varies. This suggests that caution is needed when implementing school-based suicide prevention programmes, as interventions may introduce some children to suicide for the first time. Consequently, prevention programmes should be developmentally appropriate, carefully designed, and tailored to children’s existing levels of understanding. Participants therefore emphasised teaching about broader themes, including emotional literacy, help-seeking, coping skills, and identifying trusted adults, while ensuring that suicide-related content is introduced sensitively.

Parents and professionals highlighted that suicide had become a topic of playground conversation, with schools frequently responding to disclosures of suicidal thoughts to peers, despite the topic not having been formally discussed in school. Furthermore, bereavement by suicide is becoming increasingly common (44), with professionals emphasising that there are many children who have been bereaved by, and exposed to, suicide (45). This suggests that children may already be accessing suicide-related information, often without adult oversight, increasing risk of misunderstanding and misinformation. Developmentally appropriate prevention programmes could, therefore, provide a structured and supportive context for discussing these issues. This finding supports evidence that children often disclose distressing thoughts to friends before seeking help from adults (46). A systematic review highlighted that 24% of studies showed that adolescents preferred informal support from friends and family rather than from health professionals (47), highlighting the importance of providing children with accurate information and signposting. This was particularly emphasised in relation to social media and Internet use, with many children accessing the Internet without supervision and being exposed to inappropriate and harmful content, including suicide and self-harm (48). Indeed, one in four children aged eight have their own mobile phones in the UK, and 81% of girls aged 7-21 experience upsetting behaviours online, including suicide and self-harm content (49,50), emphasising the need for online safety sessions.

Additionally, during a focus group, a primary school student suggested that suicide should be a topic to be discussed within the potential programme. While this shows that children are already aware of suicide, it also suggests an interest in learning more. This was also apparent among secondary school students, as they stated that they had learned about suicide in primary school without anyone teaching them. Primary school students expressed a further interest in mental health lessons, particularly because it was lacking in their current school. Providing CYP with factual information is important, as over-simplifying topics of suicidality can result in misinterpretation, and potentially negative outcomes (51).

In particular, the transition from primary to secondary school was highlighted as a key issue affecting CYP, and where suicide prevention education may be beneficial. This period often coincides with physical and hormonal changes (52), alongside changes in environment, routine, and academic responsibility (54,55), which may affect emotional wellbeing (57). Participants emphasised the value of accessible, school-based support (37,38), including education for primary school students on self-help, advocating for themselves and others, and knowing how and where to seek help. Although not all CYP experience mental health difficulties during the transition phase, participants felt it was still necessary to equip everyone with the knowledge and guidance on how to seek support and support others, for present or future use. This is particularly relevant to suicide prevention, given that 20% of CYP had reported that in 2023, they had seriously considered suicide according to the Youth Risk Survey (56).

However, not all participants felt that suicide prevention was appropriate in primary schools, and there were some concerns that it may be distressing. With suicide being a sensitive topic, there is a lot that needs to be considered when developing a new programme, particularly regarding the comfort and safety of the students, and how it should be integrated. Parental concerns were discussed in almost all interviews and focus groups, with the main concern being their lack of understanding of the topic and fear of the unknown. Indeed, suicide is still a stigmatised issue, and it is this stigma that induces the increased rates of suicide and self-harm (57,58), particularly because of the lack of knowledge people have on the matter (59). Whilst parental barriers were highlighted as a key issue, participants suggested that if parents were more included and informed on the subject, it would help them understanded the necessity of programme and feel more comfortable (60).

Staff comfort and confidence was another barrier, with limited training regarding suicide (61). Teachers also reported a lack of support when delivering PSHE lessons, including no supervision or debriefing opportunities. Participants had mixed opinions on whether a suicide prevention programme would be best taught by the classroom teacher or by a mental health professional due to the comfortability of the students and the knowledge of the teachers. Although, extended literature shows that there is a precedent for external professionals delivering a sessions as well as providing training for school teachers (62,63), so that they are able to support the students further. Participants also emphasised that students should be able to choose whether to attend such sessions, highlighting the importance of opt-out options to promote autonomy and allow children to seek support at their own pace (64).

Overall, given the prevalence of suicidality among CYP, participants perceived a need for a suicide prevention programme in primary schools, delivered by teachers or external professionals, instead of CYP encountering suicide-related information informally through peers or online. However, the development and possible implementation of a programme need to be carefully considered. Given the sensitivity of the subject, participants suggested that it may be most appropriate for Year 6 students (10-11 years), with a focus on self-help, available resources and techniques, and signposting for themselves or others. This could build upon lessons in earlier Years around emotional wellbeing and mental health. Online safety, including appropriate Internet use and reporting concerns, and preparation for the transition to secondary school were also key topics that participants suggested. However, participants emphasised that consent should be sought, and that the topic of suicide should be introduced sensitively, safely, and, in an age-appropriate way.

### Limitations

Although this current study explored the need for a suicide prevention programme in primary schools with a wide variety of stakeholders, some limitations should be considered. All participating schools were from Northwest England, and only one primary school contributed, which may affect the transferability of the findings. Whilst the current study had a large sample size with a variety of stakeholders, data on ethnicity and socioeconomical backgrounds as well as the gender of students was not collected, therefore it is recommended that full demographical information is collected from all participants in order to achieve representative data for future studies.

### Conclusion

Overall, the findings suggest that primary school suicide prevention education is both needed and acceptable to key stakeholders, given growing concerns about suicidality among younger children and evidence that suicide-related discussions are already occurring within peer groups. However, implementation should be approached cautiously and supported by clear guidance to ensure content is developmentally appropriate, evidence-based, and delivered safely within a robust safeguarding framework. Future policy should look to support the integration of suicide prevention within wider mental health education, alongside staff training, parental engagement, clear referral pathways, and a focus on emotional literacy, help-seeking, and access to trusted sources of support.

## Data Availability

Our data cannot be publicly shared for ethical reasons, but are available upon reasonable request from the Open Research Team at Liverpool John Moores University. Access can be requested by contacting, or by following the link: https://doi.org/10.24377/LJMU.d.00000290. When submitting your request, please detail why you want access, the project you are working on if applicable, your name and institution, and contact details. Thank you.

## Data availability statement

The data that support the findings of this study are available on request from the corresponding author. The data are not publicly available due to privacy and ethical restrictions.

## Authors Contributions

BA: Data curation, Formal Analysis, Investigation, Project Administration, Writing - Original Draft Preparation, Writing – Review & Editing.

PS: Funding Acquisition, Formal Analysis, Methodology, Supervision, Writing – Review & Editing.

MM: Methodology, Validation, Writing – Review & Editing

NS: Methodology, Validation, Writing – Review & Editing

EA: Conceptualisation, Funding Acquisition, Formal Analysis, Methodology, Supervision, Writing – Review & Editing.

## Ethical Approval

Provided by the Liverpool John Moores University Research Ethics Committee (ref: 260209LJMUREC357).

## References

1. Townsend ML, Barr KR, Miller CE, Sanzone G. Self-harm and suicidal behaviors in children: perspectives of mental health clinicians. J Pediatr Psychol. 2024 Jun 14;jsae044. doi:10.1093/jpepsy/jsae044

2. Evans R, Hurrell C. The role of schools in children and young people’s self-harm and suicide: systematic review and meta-ethnography of qualitative research. BMC Public Health. 2016 May 14;16(1):401. doi:10.1186/s12889-016-3065-2

3. Risk factors for suicide in children and young people in England - Office for National Statistics [Internet]. [cited 2026 Aug 10]. Available from: https://www.ons.gov.uk/peoplepopulationandcommunity/healthandsocialcare/mentalhealth/articles/riskfactorsforsuicideinchildrenandyoungpeopleinengland/2025-02-27

4. Programme N. Child death data release 2025. National Child Mortality Database [Internet]. 2025 Nov 13 [cited 2026 Sep 16]. Available from: https://www.ncmd.info/publications/child-death-review-data-release-2025/

5. Wise J. Children as young as 6 are increasingly attending A&amp;E in mental health crisis, analysis shows. BMJ. 2026 Sep 2;394:e100742. doi:10.1136/bmj-2026-100742 PubMed PMID: 42686360.

6. Cha CB, Franz PJ, Guzmán EM, Glenn CR, Kleiman EM, Nock MK. Suicide among Youth: Epidemiology, (Potential) Etiology, and Treatment. J Child Psychol Psychiatry. 2018 Apr;59(4):460–82. doi:10.1111/jcpp.12831 PubMed PMID: 29090457; PubMed Central PMCID: PMC5867204.

7. Bilsen J. Suicide and Youth: Risk Factors. Front Psychiatry. 2018 Oct 30;9. doi:10.3389/fpsyt.2018.00540

8. Valentine AZ, Hall SS, Sayal K, Hall CL. Waiting-list interventions for children and young people using child and adolescent mental health services: a systematic review. BMJ Ment Health. 2024 Feb 1;27(1):e300844. doi:10.1136/bmjment-2023-300844 PubMed PMID: 38302408; PubMed Central PMCID: PMC10836350.

9. Crisis in children and young people’s mental healthcare [Internet]. [cited 2026 Sep 16]. Available from: https://www.bacp.co.uk/news/news-from-bacp/2024/15-march-crisis-in-children-and-young-people-s-mental-healthcare/

10. Geoffroy MC, Bouchard S, Per M, Khoury B, Chartrand E, Renaud J, et al. Prevalence of suicidal ideation and self-harm behaviours in children aged 12 years and younger: a systematic review and meta-analysis. Lancet Psychiatry. 2022 Sep 1;9(9):703–14. doi:10.1016/S2215-0366(22)00193-6

11. Silvén Hagström A, Forinder U, Hovén E. Losing a parent to suicide: Posttraumatic stress, sense of coherence and family functioning in children, adolescents and remaining parents before attending a grief support program. Death Stud. 2025 Aug 9;49(7):841–9. doi:10.1080/07481187.2024.2361759 PubMed PMID: 38843028.

12. Rodway C, Ibrahim S, Tham SG, Turnbull P, Kapur N, Appleby L. Bereavement and suicide bereavement as an antecedent of suicide in children and young people: Prevalence and characteristics. J Affect Disord. 2022 Mar;300:280–8. doi:10.1016/j.jad.2021.12.063

13. Cerel J, Maple M, van de Venne J, Brown M, Moore M, Flaherty C. Suicide Exposure in the Population: Perceptions of Impact and Closeness. Suicide Life Threat Behav. 2017;47(6):696– 708. doi:10.1111/sltb.12333

14. Skei-Larssen E, Grimholt TK, Qin P. Risk of deliberate self-harm and suicide in children bereaved from parental death by specific natural and external causes – A national study. J Affect Disord. 2026 Mar;397:120984. doi:10.1016/j.jad.2025.120984

15. Wardley A. CHURCHILL FELLOWSHIP REPORT.

16. Harris J, Nowland R, Todd M. Intersecting Identities and Adjustment to the Primary-Secondary School Transition: An Integrative Review. J Educ Res Pract. 2024 Feb 10;14(1). doi:10.5590/JERAP.2024.14.1.05

17. Mumford J, Birchwood J. Transition: a systematic review of literature exploring the experiences of pupils moving from primary to secondary school in the UK. Pastor Care Educ. 2021 Oct 2;39(4):377–400. doi:10.1080/02643944.2020.1855670

18. Dimitrellou E, Koutsouris G, Pearson A, Yildirim H. Transitioning in early adolescence: rethinking the role of socioemotional learning (SEL) in the UK school curriculum to foster adaptive coping. Pastor Care Educ. 2025 Apr 29;0(0):1–21. doi:10.1080/02643944.2025.2495664

19. A Theoretically Grounded Exploration of the Social and Emotional Outcomes of Transition to Secondary School [Internet]. [cited 2026 Aug 10]. Available from: https://journals.sagepub.com/doi/epub/10.1017/jgc.2012.26 doi:10.1017/jgc.2012.26

20. Harwood S, Eaves S. Conceptualising technology, its development and future: The six genres of technology. Technol Forecast Soc Change. 2020 Nov;160:120174. doi:10.1016/j.techfore.2020.120174 PubMed PMID: 32904525; PubMed Central PMCID: PMC7456399.

21. Borges D, Pinto I, Santos O, Moura I, Ferreira IR, Macedo AP, et al. On–Off Childhood? A Rapid Review of the Impact of Technology on Children’s Health. Healthcare. 2025 Jul 21;13(14):1769. doi:10.3390/healthcare13141769 PubMed PMID: 40724793; PubMed Central PMCID: PMC12295127.

22. Wang J, Iannotti RJ, Luk JW. Computer Use and Internet Bullying among US Adolescents: Gender and Grade Differences. J Ehealth Technol Appl. 2009 Dec;7(2):72–7. PubMed PMID: 26413176; PubMed Central PMCID: PMC4583206.

23. Prkno D, Grafe N, Schulz MS, Kiess W, Poulain T. Children’s and adolescents’ negative internet experiences and the association with quality of life and behavioural difficulties: a cross-sectional study. BMJ Paediatr Open. 2025 Apr 17;9(1):e003135. doi:10.1136/bmjpo-2024-003135 PubMed PMID: 40246585; PubMed Central PMCID: PMC12007062.

24. Muppalla SK, Vuppalapati S, Reddy Pulliahgaru A, Sreenivasulu H. Effects of Excessive Screen Time on Child Development: An Updated Review and Strategies for Management. Cureus. 15(6):e40608. doi:10.7759/cureus.40608 PubMed PMID: 37476119; PubMed Central PMCID: PMC10353947.

25. Robinson J, La Sala L, Kenny B, Cooper C, Lamblin M, Spittal M, et al. How do Australian social media users experience self-harm and suicide-related content? A National cross-sectional survey comparing young people and adults. BMC Public Health. 2025 Dec 4;26(1):384. doi:10.1186/s12889-025-25646-0

26. Programme of Study for PSHE education (KS1-5) [Internet]. [cited 2026 Sep 30]. Available from: https://pshe-association.org.uk/guidance/ks1-5/programme-of-study

27. Hayes D, Moore A, Stapley E, Humphrey N, Mansfield R, Santos J, et al. Promoting mental health and wellbeing in schools: examining Mindfulness, Relaxation and Strategies for Safety and Wellbeing in English primary and secondary schools: study protocol for a multi-school, cluster randomised controlled trial (INSPIRE). Trials. 2019 Nov 21;20(1):640. doi:10.1186/s13063-019-3762-0

28. Bem D, Connor C, Palmer C, Channa S, Birchwood M. Frequency and preventative interventions for non-suicidal self-injury and suicidal behaviour in primary school-age children: a scoping review protocol [Internet]. 2017 Jul 1. doi:10.1136/bmjopen-2017-017291

29. Breaking The Silence. Grassroots Suicide Prevention [Internet]. [cited 2026 Aug 10]. Available from: https://prevent-suicide.org.uk/breaking-the-silence/

30. Schilling EA, Aseltine RH, James A. The SOS Suicide Prevention Program: Further Evidence of Efficacy and Effectiveness. Prev Sci. 2016 Feb 1;17(2):157–66. doi:10.1007/s11121-015-0594-3

31. Pokharel Poudel R, Perumbil Pathrose S, Jefferies D, Ramjan LM. Effectiveness of Suicide Prevention Programmes Among Adolescents and Sociocultural Adaptation of Programmes: A Systematic Review. Int J Ment Health Nurs. 2025;34(2):e70038. doi:10.1111/inm.70038

32. Ashworth E, Thompson J, Saini P. “It’s like an epidemic, we don’t know what to do”: The perceived need for and benefits of a suicide prevention programme in UK schools. Br J Educ Psychol. 2024 Sep 1;94(3):824–38. doi:10.1111/bjep.12683

33. Hennefield L, Luking KR, Tillman R, Barch DM, Luby JL, Thompson RJ. Asking Preadolescents About Suicide Is Not Associated With Increased Suicidal Thoughts. J Am Acad Child Adolesc Psychiatry. 2026 Jan;65(1):34–41. doi:10.1016/j.jaac.2025.03.025 PubMed PMID: 40188866; PubMed Central PMCID: PMC12866979.

34. Walsh EH, McMahon J, Herring MP. Research Review: The effect of school-based suicide prevention on suicidal ideation and suicide attempts and the role of intervention and contextual factors among adolescents: a meta-analysis and meta-regression. J Child Psychol Psychiatry. 2022 Aug;63(8):836–45. doi:10.1111/jcpp.13598 PubMed PMID: 35289410; PubMed Central PMCID: PMC9544521.

35. Nadeem E, Santiago CD, Kataoka SH, Chang VY, Stein BD. School personnel experiences in notifying parents about their child’s risk for suicide: lessons learned. J Sch Health. 2016 Jan;86(1):3–10. doi:10.1111/josh.12346 PubMed PMID: 26645415; PubMed Central PMCID: PMC4747656.

36. GOV.UK [Internet]. 2026 [cited 2026 Sep 28]. Keeping children safe in education. Available from: https://www.gov.uk/government/publications/keeping-children-safe-in-education--2

37. Calear AL, Christensen H. Systematic review of school-based prevention and early intervention programs for depression. J Adolesc. 2010 Jun 1;33(3):429–38. doi:10.1016/j.adolescence.2009.07.004

38. McCarthy M, Wynne S, Ashworth E, Hendriks O, Saini P. Behind the data: Reflections from conducting school-based suicide prevention research with young people in the United Kingdom. Montague-Cardoso K, editor. PLOS Ment Health. 2025 Oct 30;2(10):e0000476. doi:10.1371/journal.pmen.0000476

39. Ayer L, Stevens C, Reider E, Sims B, Colpe L, Pearson J. Preventing Youth Suicide: Potential “Crossover Effects” of Existing School-Based Programs. Prev Sci. 2023 Feb 1;24(2):382–92. doi:10.1007/s11121-022-01473-2

40. Ali MM, West K, Teich JL, Lynch S, Mutter R, Dubenitz J. Utilization of Mental Health Services in Educational Setting by Adolescents in the United States. J Sch Health. 2019;89(5):393–401. doi:10.1111/josh.12753

41. Graziano F, Davico C, Giordano I, Lonardelli E, Marcotulli D, Calandri E. Teachers as Gatekeepers in Adolescent Suicide Prevention: The Role of Suicide-Related Knowledge, Empathy, and Collaborative Self-Efficacy. Behav Sci. 2026 Mar;16(3):409. doi:10.3390/bs16030409

42. Ritchie J, Spencer L. Qualitative data analysis for applied policy research. In: Analyzing Qualitative Data. Routledge; 1994.

43. Miller AB, Sheridan MA, Pelletier-Baldelli A, Sirhal SR, Lu J, Boda SS, et al. Suicidal Thoughts and Behaviors in Elementary School–Aged Youth: A Pilot Study in 5- to 10-Year-Olds. JAACAP Open. 2025 Sep 1;3(3):527–37. doi:10.1016/j.jaacop.2025.01.006

44. McDonnell S, Flynn S, Shaw J, Smith S, McGale B, Hunt IM. Suicide bereavement in the UK: Descriptive findings from a national survey. Suicide Life Threat Behav. 2022 Oct;52(5):887–97. doi:10.1111/sltb.12874 PubMed PMID: 35611626; PubMed Central PMCID: PMC9790485.

45. Childhood Bereavement Network [Internet]. [cited 2026 Sep 16]. Key statistics. Available from: https://childhoodbereavementnetwork.org.uk/about/media-centre/evidence/key-statistics

46. McGuire K, London K. A retrospective approach to examining child abuse disclosure. Child Abuse Negl. 2020 Jan 1;99:104263. doi:10.1016/j.chiabu.2019.104263

47. Radez J, Reardon T, Creswell C, Lawrence PJ, Evdoka-Burton G, Waite P. Why do children and adolescents (not) seek and access professional help for their mental health problems? A systematic review of quantitative and qualitative studies. Eur Child Adolesc Psychiatry. 2021;30(2):183–211. doi:10.1007/s00787-019-01469-4 PubMed PMID: 31965309; PubMed Central PMCID: PMC7932953.

48. Hernandez JM, Ben-Joseph EP, Reich S, Charmaraman L. Parental Monitoring of Early Adolescent Social Technology Use in the US: A Mixed-Method Study. J Child Fam Stud. 2024;33(3):759–76. doi:10.1007/s10826-023-02734-6 PubMed PMID: 40621594; PubMed Central PMCID: PMC12227363.

49. Screen time: impacts on education and wellbeing.

50. Girlguiding [Internet]. [cited 2026 Sep 16]. Girls’ Attitudes Survey. Available from: https://www.girlguiding.org.uk/girlsattitudes/

51. Understanding Suicide. Grassroots Suicide Prevention [Internet]. [cited 2026 Sep 16]. Available from: https://prevent-suicide.org.uk/understanding-suicide/

52. Alloy LB, Hamilton JL, Hamlat EJ, Abramson LY. Pubertal Development, Emotion Regulatory Styles, and the Emergence of Sex Differences in Internalizing Disorders and Symptoms in Adolescence. Clin Psychol Sci J Assoc Psychol Sci. 2016 Sep;4(5):867–81. doi:10.1177/2167702616643008 PubMed PMID: 27747141; PubMed Central PMCID: PMC5061504.

53. Bagnall CL. Talking about School Transition (TaST): an emotional centred intervention to support children over primary-secondary school transition. Pastor Care Educ. 2020 Apr 2;38(2):116–37. doi:10.1080/02643944.2020.1713870

54. Beatson R, Quach J, Canterford L, Farrow P, Bagnall C, Hockey P, et al. Improving primary to secondary school transitions: A systematic review of school-based interventions to prepare and support student social-emotional and educational outcomes. Educ Res Rev. 2023 Aug;40:100553. doi:10.1016/j.edurev.2023.100553

55. Spernes K. The transition between primary and secondary school: a thematic review emphasising social and emotional issues. Res Pap Educ. 2022 May 4;37(3):303–20. doi:10.1080/02671522.2020.1849366

56. CDC. Youth Risk Behavior Surveillance System (YRBSS) [Internet]. 2025 [cited 2026 Oct 5]. YRBS Data Summary & Trends Report. Available from: https://www.cdc.gov/yrbs/dstr/index.html

57. Costanza A, Favre S, Richard-Lepouriel H. Suicidality and mental health stigma and their impact on suicidal ideation disclosure: A narrative review. J Affect Disord Rep. 2026 Apr 1;24:101054. doi:10.1016/j.jadr.2026.101054

58. Wyllie JM, Robb KA, Belkadi N, O’Connor RC. “I still think there’s like a huge stigma”: Understanding suicide-related stigma among those with lived experiences. Death Stud. 2025 Sep 8;0(0):1–13. doi:10.1080/07481187.2025.2556106 PubMed PMID: 40920066.

59. Kågström A, Guerrero Z, Aliev AA, Tomášková H, Rüsch N, Ouali U, et al. Mental health stigma and its consequences: a systematic scoping review of pathways to discrimination and adverse outcomes. eClinicalMedicine. 2025 Oct 23;89:103588. doi:10.1016/j.eclinm.2025.103588 PubMed PMID: 41210382; PubMed Central PMCID: PMC12593596.

60. ResearchGate [Internet]. [cited 2026 Sep 16]. The Importance of Cooperation between Parents and School. Available from: https://www.researchgate.net/publication/393379190_The_Importance_of_Cooperation_between_Parents_and_School

61. Lawson GM, Orapallo A, Ginsburg GS, Brewton A, Baker CN, Azad G. Teacher-Delivered Mental Health Interventions: Promises, Challenges, and Recommendations for Future Directions. Adm Policy Ment Health Ment Health Serv Res. 2025 Nov 1;52(6):1296–310. doi:10.1007/s10488-025-01467-6

62. Ashworth E, Hanlon C, McCarthy M, Hunt A, Wynne S, Foster R, et al. Multi-modal approach to preventing suicide in schools: a regionally-based UK pilot study. Front Child Adolesc Psychiatry. 2026 Mar 5;5. doi:10.3389/frcha.2026.1768052

63. Lindow JC, Hughes JL, South C, Minhajuddin A, Gutierrez L, Bannister E, et al. The Youth Aware of Mental Health (YAM) Intervention: Impact on Help-Seeking, Mental Health Knowledge, and Stigma in US Adolescents. J Adolesc Health Off Publ Soc Adolesc Med. 2020 Jul;67(1):101–7. doi:10.1016/j.jadohealth.2020.01.006 PubMed PMID: 32115325; PubMed Central PMCID: PMC7311230.

64. de Soet R, Vermeiren RRJM, Bansema CH, Nijland L, Mulder EA, Nooteboom LA. Balancing autonomy and safety in care for youth with severe and enduring mental health problems. Eur Child Adolesc Psychiatry. 2026 Apr 1;35(4):1211–22. doi:10.1007/s00787-025-02930-3

